# Artificial intelligence-enhanced risk stratification of cancer therapeutics-related cardiac dysfunction using electrocardiographic images

**DOI:** 10.1101/2024.03.12.24304047

**Authors:** Evangelos K. Oikonomou, Veer Sangha, Lovedeep S. Dhingra, Arya Aminorroaya, Andreas Coppi, Harlan M. Krumholz, Lauren A. Baldassarre, Rohan Khera

## Abstract

**Background:** Risk stratification strategies for cancer therapeutics-related cardiac dysfunction (CTRCD) rely on serial monitoring by specialized imaging, limiting their scalability.

**Objectives:** To examine an artificial intelligence (AI)-enhanced electrocardiographic (AI-ECG) surrogate for imaging risk biomarkers, and its association with CTRCD.

**Methods:** Across a five-hospital U.S.-based health system (2013-2023), we identified patients with breast cancer or non-Hodgkin lymphoma (NHL) who received anthracyclines (AC) and/or trastuzumab (TZM), and a control cohort receiving immune checkpoint inhibitors (ICI). We deployed a validated AI model of left ventricular systolic dysfunction (LVSD) to ECG images (≥0.1, positive screen) and explored its association with i) global longitudinal strain (GLS) measured within 15 days (*n*=7,271 pairs); ii) future CTRCD (new cardiomyopathy, heart failure, or left ventricular ejection fraction [LVEF]<50%), and LVEF<40%. In the ICI cohort we correlated baseline AI-ECG-LVSD predictions with downstream myocarditis.

**Results:** Higher AI-ECG LVSD predictions were associated with worse GLS (−18% [IQR:-20 to −17%] for predictions<0.1, to −12% [IQR:-15 to −9%] for ≥0.5 (*p*<0.001)). In 1,308 patients receiving AC/TZM (age 59 [IQR:49-67] years, 999 [76.4%] women, 80 [IQR:42-115] follow-up months) a positive baseline AI-ECG LVSD screen was associated with ∼2-fold and ∼4.8-fold increase in the incidence of the composite CTRCD endpoint (adj.HR 2.22 [95%CI:1.63-3.02]), and LVEF<40% (adj.HR 4.76 [95%CI:2.62-8.66]), respectively. Among 2,056 patients receiving ICI (age 65 [IQR:57-73] years, 913 [44.4%] women, follow-up 63 [IQR:28-99] months) AI-ECG predictions were not associated with ICI myocarditis (adj.HR 1.36 [95%CI:0.47-3.93]).

**Conclusion:** AI applied to baseline ECG images can stratify the risk of CTRCD associated with anthracycline or trastuzumab exposure.

**CONDENSED ABSTRACT:** There is an unmet need for scalable and affordable biomarkers to stratify the risk of cancer therapeutics-related cardiac dysfunction (CTRCD). In this hospital system-based, decade-long cohort of patients without cardiomyopathy receiving anthracyclines or trastuzumab, a validated artificial intelligence algorithm applied to baseline electrocardiographic (AI-ECG) images identified individuals with a 2-fold and 4.8-fold risk of developing any cardiomyopathy or left ventricular ejection fraction <40%, respectively. This supports a role for AI-ECG interpretation of images as a scalable approach for the baseline risk stratification of patients initiating cardiotoxic chemotherapy.

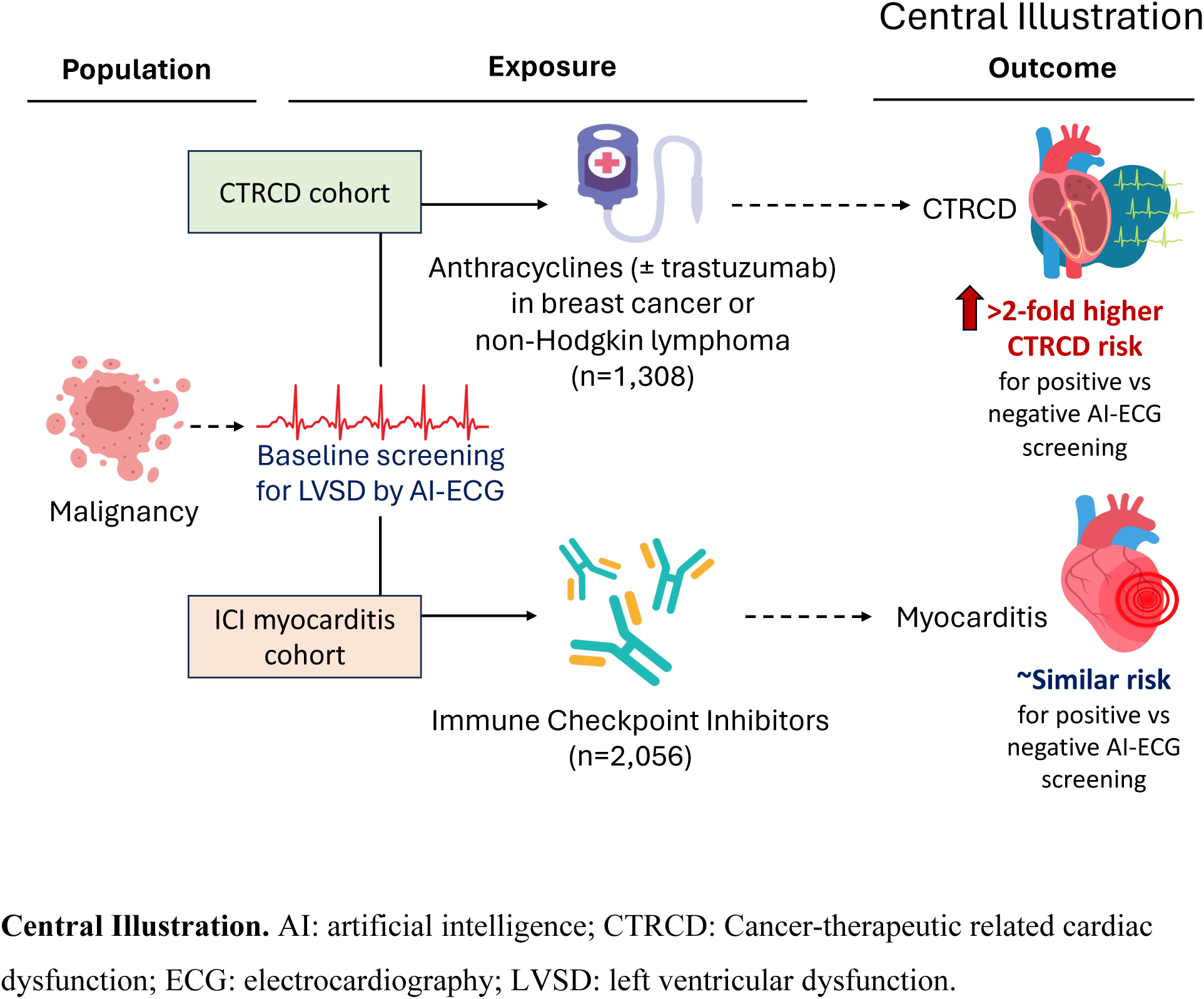

## INTRODUCTION

Several oncological therapies that have enabled improvements in cancer-specific survival pose long-term deleterious effects on the cardiovascular system.^3^ There is an emerging need to design scalable and affordable protocols for the efficient risk stratification of cardiac therapeutics-related cardiac dysfunction (CTRCD), predominantly for patients exposed to widely used chemotherapy, targeted agents, and immunotherapy, such as anthracyclines,^4^ human epidermal growth factor receptor-2 (HER2) receptor inhibitors, and immune checkpoint inhibitors (ICI).^5–7^ Recent guideline statements have summarized a growing body of literature on the appropriate risk stratification, prevention, and management of comorbid or incident cardiovascular disease,^3, 8, 9^ boosted by the development in non-invasive biomarkers,^10^ multimodal imaging,^11^ and, more recently, artificial intelligence (AI).^12–15^

Despite this, the accurate identification of high-risk individuals *before* the initiation of therapy remains an elusive goal. A recent systematic review concluded that there is currently insufficient evidence for personalized risk prediction tools in this population.^16^ However, trials of screening strategies using advanced echocardiographic indices, such as global longitudinal strain (GLS)^17^ have suggested possible benefit from flagging subclinical left ventricular systolic dysfunction and guiding the deployment of potentially disease-modifying therapies.^18–25^ Therefore, there is an unmet need to design scalable yet personalized tools to enable the timely identification of patients who might benefit from more intensive follow-up. Furthermore, it is critical that these tools are accessible and affordable, ensuring equitable and broad access across high and low-resource settings.

Building on recent advances in AI-ECG (AI-electrocardiography), a recently developed and validated deep learning model trained against concurrent echocardiographic labels has been shown to predict both the presence and future risk of left ventricular systolic dysfunction (LVSD) directly from ECG images.^26, 27^ This tool, accessible to the end-user on any browser or smartphone interface enables clinicians to screen for LVSD at the point-of-care. In the present study, we first examine the cross-sectional correlation between AI-ECG LVSD predictions and concurrent GLS measurements on transthoracic echocardiography. Next, we hypothesized that AI applied to ECG images of patients undergoing potential cardiotoxic chemotherapy with anthracyclines or trastuzumab may identify individuals at higher risk of CTRCD independent of traditional cardiovascular factors and in the absence of clinical LVSD at the time of treatment initiation. We evaluate the specificity of this association of AI-ECG for future LVSD risk using a parallel analysis of patients on ICI therapy followed for the incidence of myocarditis, an immune-related adverse event with distinct underlying pathophysiology.^5, 6^

## METHODS

### Data Source and Patient Population

This was a retrospective cohort study of patients who received treatment with anthracyclines, trastuzumab, or ICI between January 1, 2013, and December 5, 2023. We identified all individuals within the electronic health record (EHR) of the Yale-New Haven Health (YNHH) system, a system spanning five hospitals and several clinics across Connecticut and Rhode Island for eligible individuals who were 18 years or older at the time of their treatment to define two distinct cohorts. Our main cohort consisted of adult patients with breast cancer or non-Hodgkin lymphoma who received treatment with anthracycline (doxorubicin, daunorubicin, valrubicin, epirubicin, idarubicin or mitoxantrone), and/or trastuzumab (breast cancer) within five years of their index cancer diagnosis, followed for the primary outcome of CTRCD. For a negative control analysis, we further identified a broad population of adult patients with various forms of cancer undergoing immunotherapy with any combination of ICI (pembrolizumab, ipilimumab, nivolumab, atezolizumab, cemiplimab, avelumab, durvalumab, teremelimumab, relatlimab, toripalimab) who were followed for the primary outcome of myocarditis (**Central Illustration**).

To be eligible for inclusion in our analysis, we required the presence of a 12-lead ECG within 36 months before, up to 1 month after treatment initiation. We excluded cases where patients received any of these therapies in the context of a clinical trial, as well as patients who had a history of LVEF <50% and/or a diagnosis of cardiomyopathy or heart failure at baseline. For individuals with two or more cancers, we defined the chronologically first cancer diagnosis and treatment regimen as the index diagnosis and exposure for that participant. Furthermore, to prevent data leakage from our model’s training, we excluded any ECGs that were previously used in the training of the image-based AI-ECG model.^26^

### Study Covariates

We collected information on demographics (age, sex, race, ethnicity), medications, and diagnoses from the electronic health record (EHR) of the hospital system. We defined all diagnoses, inclusive of cancer type, as well as traditional cardiovascular risk factors and comorbidities (i.e., hypertension, diabetes mellitus, chronic kidney disease, peripheral arterial disease, stroke) using a combination of ICD-9-CM and ICD-10-CM (International Classification of Diseases [ICD] 9 and 10 Clinical Modification) diagnosis codes. ICD codes were mapped to categories defined using CCSR (Clinical Classification Software - Refined) codes, curated and maintained by the Agency for Healthcare Research and Quality (**Online Table 1**). Where applicable, diagnoses of ischemic heart disease, and peripheral arterial disease were supplemented by current procedural terminology (CPT) codes describing disease-defining procedures, such as percutaneous coronary intervention (PCI), coronary artery bypass grafting (CABG) or carotid or peripheral arterial interventions (**Online Table 2**). To account for possible delays in administrative claims and recordings, baseline procedure and diagnosis code recorded up before and up to 30 days after treatment initiation.

**Table 1.**
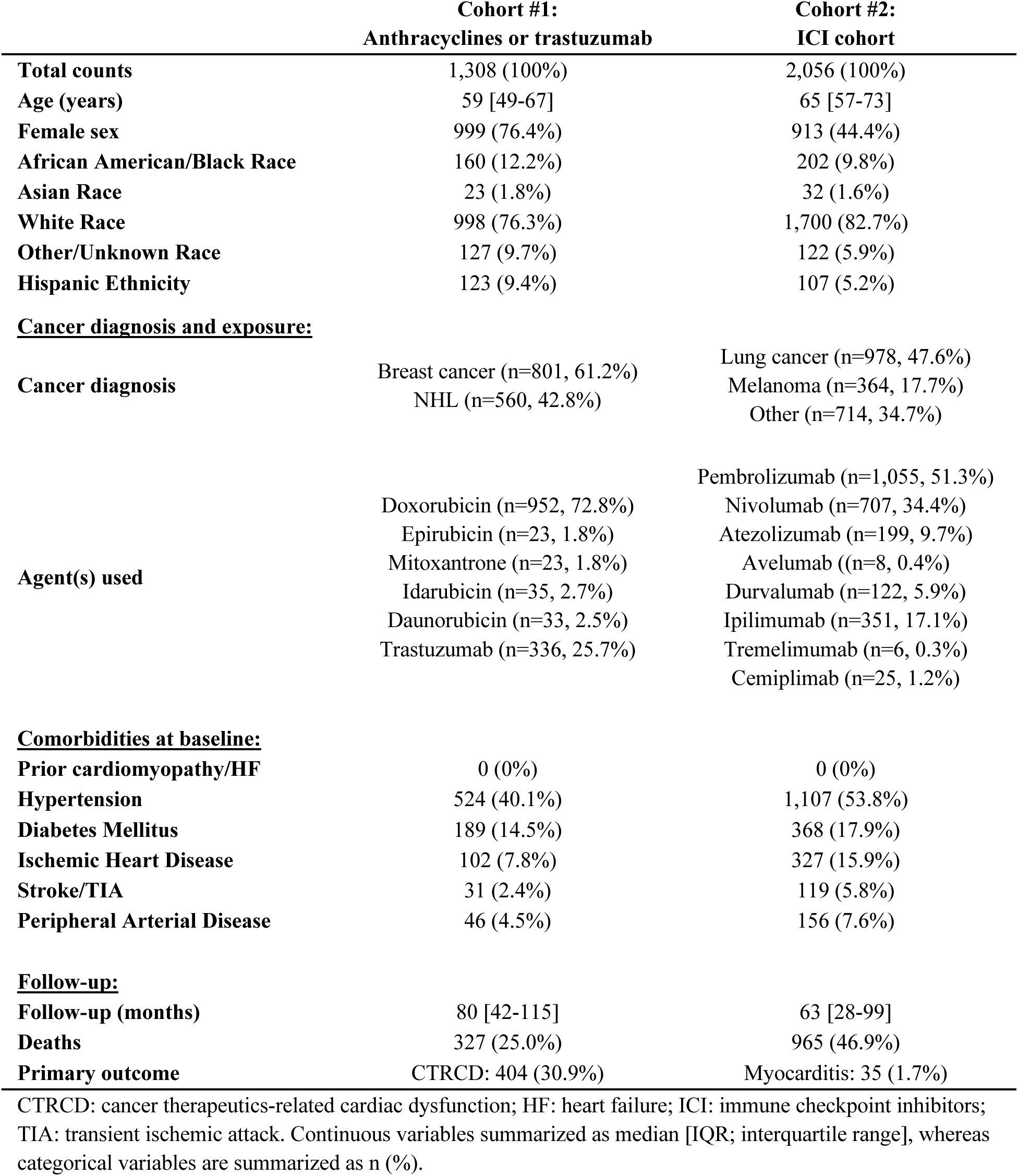
Table of demographics.

|  | **Cohort #1:**<br>**Anthracyclines or trastuzumab** | **Cohort #2:**<br>**ICI cohort** |
| --- | --- | --- |
| **Total counts** | 1,308 (100%) | 2,056 (100%) |
| **Age (years)** | 59 [49-67] | 65 [57-73] |
| **Female sex** | 999 (76.4%) | 913 (44.4%) |
| **African American/Black Race** | 160 (12.2%) | 202 (9.8%) |
| **Asian Race** | 23 (1.8%) | 32 (1.6%) |
| **White Race** | 998 (76.3%) | 1,700 (82.7%) |
| **Other/Unknown Race** | 127 (9.7%) | 122 (5.9%) |
| **Hispanic Ethnicity** | 123 (9.4%) | 107 (5.2%) |
| **<u>Cancer diagnosis and exposure:</u>** |  |  |
| **Cancer diagnosis** | Breast cancer (n=801, 61.2%)<br>NHL (n=560, 42.8%) | Lung cancer (n=978, 47.6%)<br>Melanoma (n=364, 17.7%)<br>Other (n=714, 34.7%) |
| **Agent(s) used** | Doxorubicin (n=952, 72.8%)<br>Epirubicin (n=23, 1.8%)<br>Mitoxantrone (n=23, 1.8%)<br>Idarubicin (n=35, 2.7%)<br>Daunorubicin (n=33, 2.5%)<br>Trastuzumab (n=336, 25.7%) | Pembrolizumab (n=1,055, 51.3%)<br>Nivolumab (n=707, 34.4%)<br>Atezolizumab (n=199, 9.7%)<br>Avelumab ((n=8, 0.4%)<br>Durvalumab (n=122, 5.9%)<br>Ipilimumab (n=351, 17.1%)<br>Tremelimumab (n=6, 0.3%)<br>Cemiplimab (n=25, 1.2%) |
| **<u>Comorbidities at baseline:</u>** |  |  |
| **Prior cardiomyopathy/HF** | 0 (0%) | 0 (0%) |
| **Hypertension** | 524 (40.1%) | 1,107 (53.8%) |
| **Diabetes Mellitus** | 189 (14.5%) | 368 (17.9%) |
| **Ischemic Heart Disease** | 102 (7.8%) | 327 (15.9%) |
| **Stroke/TIA** | 31 (2.4%) | 119 (5.8%) |
| **Peripheral Arterial Disease** | 46 (4.5%) | 156 (7.6%) |
| **<u>Follow-up:</u>** |  |  |
| **Follow-up (months)** | 80 [42-115] | 63 [28-99] |
| **Deaths** | 327 (25.0%) | 965 (46.9%) |
| **Primary outcome** | CTRCD: 404 (30.9%) | Myocarditis: 35 (1.7%) |

CTRCD: cancer therapeutics-related cardiac dysfunction; HF: heart failure; ICI: immune checkpoint inhibitors; TIA: transient ischemic attack. Continuous variables summarized as median [IQR; interquartile range], whereas categorical variables are summarized as n (%).

### AI-Guided Stratification Of LVSD Risk On ECG Images

Baseline 12-lead ECG studies were extracted and pre-processed using our previously described pipeline,^26^ to create images of ECGs. This approach simulates a clinical workflow where a provider may wish to directly access the algorithm through their browser or smartphone application by taking a photo or screenshot of a 12-lead ECG. We then deployed our previously fine-tuned convolutional neural network (CNN) to obtain a probability of LVSD, ranging from 0 to 1 (with a previously validated cutoff of 0.1 to discriminate positive from negative screens).^26^ An online version of the model with an interactive user interface is publicly available for research use at: https://www.cards-lab.org/ecgvision-lv.^28^

### Saliency maps

For explainability, we used the GradCAM (Gradient-weighted Class Activation)^29^ method to highlight the parts of the image that were the most important in predicting the LVSD class. This is done by calculating the gradients on the final stack of filters in the deep learning model (EfficientNet-B3) architecture for each case, performing a global average pooling of the gradients in each filter, and emphasizing those contributing to the prediction. These are then multiplied by their importance weights and combined across filters to generate GradCAM heatmaps, as previously reported.^26^ GradCAM intensities, from 0 to 1, are converted to a color range using the jet colormap, which is upsampled and overlaid to the original image size with a transparency level of alpha=0.3.

### Study Outcomes

In the imaging correlation analysis, we evaluated the cross-sectional association of AI-ECG based LVSD classification with baseline echocardiography-determined GLS (%), as reported by the cardiologist who interpreted the original study. This analysis was restricted to GLS measurements performed within 15 days (−15 to +15 days) of the respective ECG.

In the clinical outcomes analysis, we evaluated the incidence of CTRCD, which was defined using a combination of diagnosis codes and/or objectively defined left ventricular systolic dysfunction. This included the time-to-first diagnosis of a relevant cardiomyopathy (**Online Table 1**), left ventricular systolic dysfunction, or heart failure, or an echocardiogram with a post-treatment LVEF below 50%. Secondary outcomes were the time-to-first-LVEF <50%, or <40%, as determined on echocardiography. Of note, given variations in the definitions across different guidelines,^3, 9^ as well as the time period of our study, which pre-dated much of the current evidence on the role of GLS,^17, 18^ we did not include relative changes in GLS in our outcome.

In the ICI cohort, the primary outcome was the time-to-myocarditis. Given the multifactorial nature of this diagnosis, which often involves multimodality imaging and serum biomarker assessment integrated with relevant clinical history, symptoms, and clinical exam findings, we opted to define this as the first appearance of a myocarditis-specific ICD/CCS(R) code after treatment. In both cohorts, we also analyzed the time-to-all-cause mortality, as a secondary outcome. In a secondary analysis, we provided descriptive summaries of AI-ECG LVSD predictions in discrete time intervals leading up to CTRCD diagnosis (or last known follow-up for controls).

### Statistical Analysis

Continuous variables are summarized using median (interquartile range; IQR) or mean (standard deviation) values, as appropriate, and categorical variables as counts (percentages, %). For the imaging outcome analyses, the distribution of GLS across subgroups of AI-ECG predictions were summarized using density plots and compared using the non-parametric Kruskal-Wallis test.

We evaluated the AI-ECG LVSD model as a predictor for subsequent CTRCD. This was done using two approaches. First, we assessed the association between the binary screening profile (positive if score/probability of AI-ECG LVSD 0.1 or greater, negative if <0.1) with the downstream incidence of the primary and secondary outcomes in multivariable Cox regression models adjusted for the patient’s baseline age, sex, race, and ethnicity, as well as traditional cardiovascular risk factors, including the presence of hypertension, diabetes mellitus, coronary artery disease, peripheral arterial disease, preceding stroke, chronic kidney disease, and smoking. Second, to evaluate the role of AI-ECG in identifying periods leading up to CTRCD, we compared the log-transformed AI-ECG scores across 12-month intervals (48-36, 36-24, 24-12, 12-0 months before CTRCD diagnosis or last known follow-up) between patients with versus without CTRCD using two-way analysis of variance (ANOVA). This was followed by post-hoc pairwise comparisons within each time period between the CTRCD and non-CTRCD groups. These were performed by independent sample t-tests with alpha adjusted for multiple comparisons using the Bonferroni method.

Across all multivariable regression models, covariates with missing values were imputed using non-parametric chained equation imputation with random forests by including all model covariates.^30^ All statistical tests were two-sided with a significance level of 0.05. Analyses were performed using Python (version 3.11.2).

#### Ethical Approval

The Yale Institutional Review Board (IRB) approved our study and waived the requirement for informed consent for this retrospective, chart review study.

## RESULTS

### Summary of the overall patient population

Our initial chart review identified 14,506 unique patients (median age of 67 [IQR 58-76] years, 8,775 (60.5%) women) who received any of the 3 classes of medications between 2013 and 2023 (n=6,176 anthracyclines, n=2,043 trastuzumab, n=6330 ICI), with the most common cancer types including breast cancer (n=4,504), respiratory cancers (n=3,704), Non-Hodgkin lymphomas (n=2,249), and melanoma (n=1,291). The cumulative exposure to one or more classes of therapies across patients with one or more cancer types is summarized in **Figure 1**.

**Figure 1.**
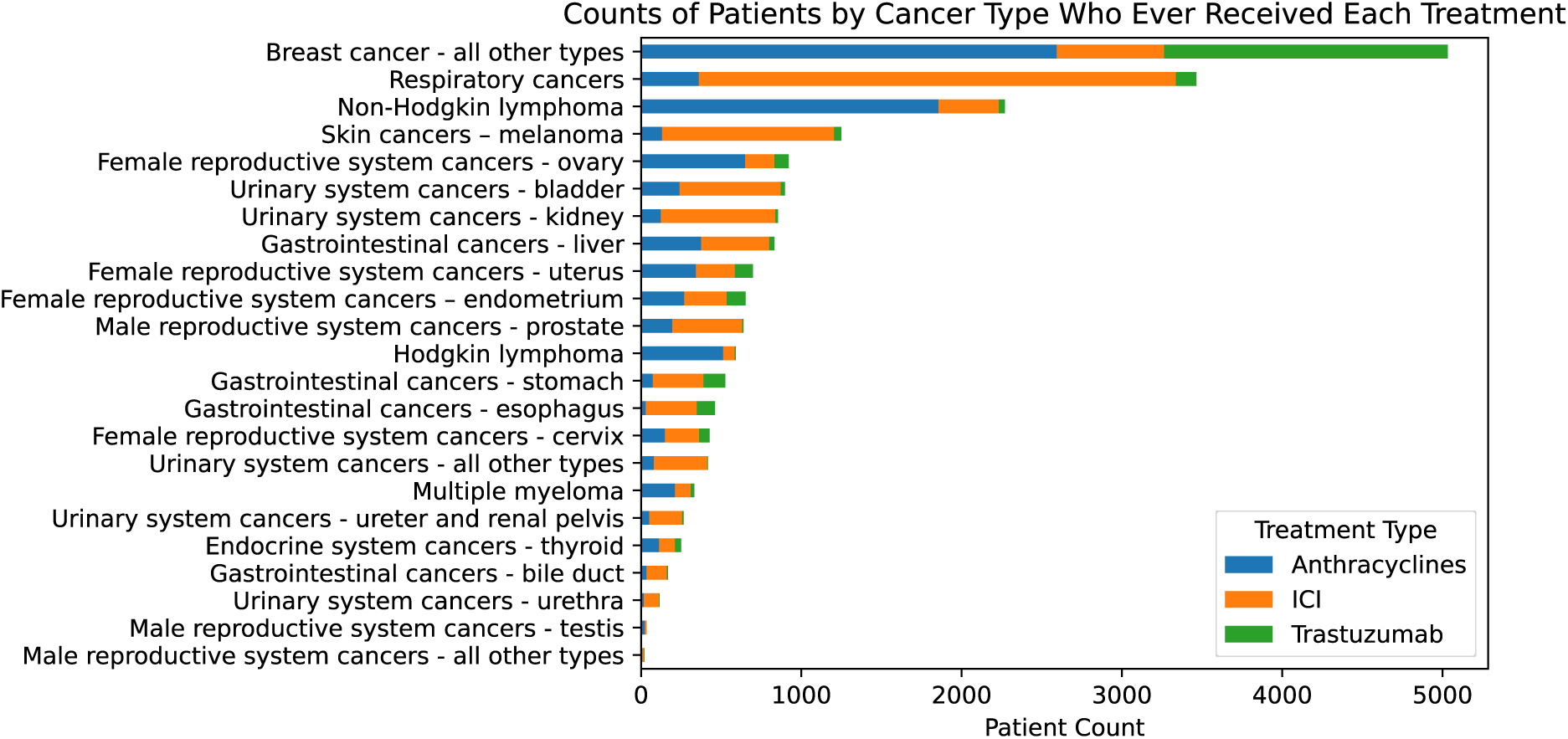
Counts of unique patients stratified by cancer types and cumulative exposure to anthracyclines, trastuzumab or ICI. Counts reflect any cancer diagnosis and any exposure during the study period. For instance, an individual participant may have had two or more cancer diagnoses and received different agents for different indication. ICI: immune checkpoint inhibitors.

### Cross-sectional correlation between AI-ECG and GLS

There were 58,876 and 53,296 ECGs identified in 6,098 and 5,277 patients receiving anthracyclines/trastuzumab or ICI, with 10,801 and 2,880 ECGs performed before or within 30 days of treatment initiation, respectively (**Figure 2**). We identified 7,271 pairs of 12-lead ECGs and transthoracic echocardiograms with GLS within 15 days of each other among 2,004 unique patients. There was a graded relationship between higher AI-ECG LVSD scores and lower GLS, indicative of worse LV systolic function, with the median [IQR] GLS ranging from −18 [-20 to - 17]% for AI-ECG scores <0.1 (n=6333), to −16 [-19 to −13]% for AI-ECG scores ≥0.1 but <0.5 (n=759), and −12 [-15 to −9]% for AI-ECG scores ≥0.5 (n=179) (*p*<0.001) (**Figure 3a**). Of note, this was consistent in a subgroup analysis that only included pairs with concurrent LVEF measurements of ≥50% (AI-ECG scores <0.1 (n=6049): GLS −19 [-20 to −17]%, 0.1 to 0.5 (n=536): GLS −17 [-20 to −15]%, and ≥0.5 (n=60): GLS −16 [-19 to −14]%, *p*<0.001) (**Figure 3b**).

**Figure 2.**
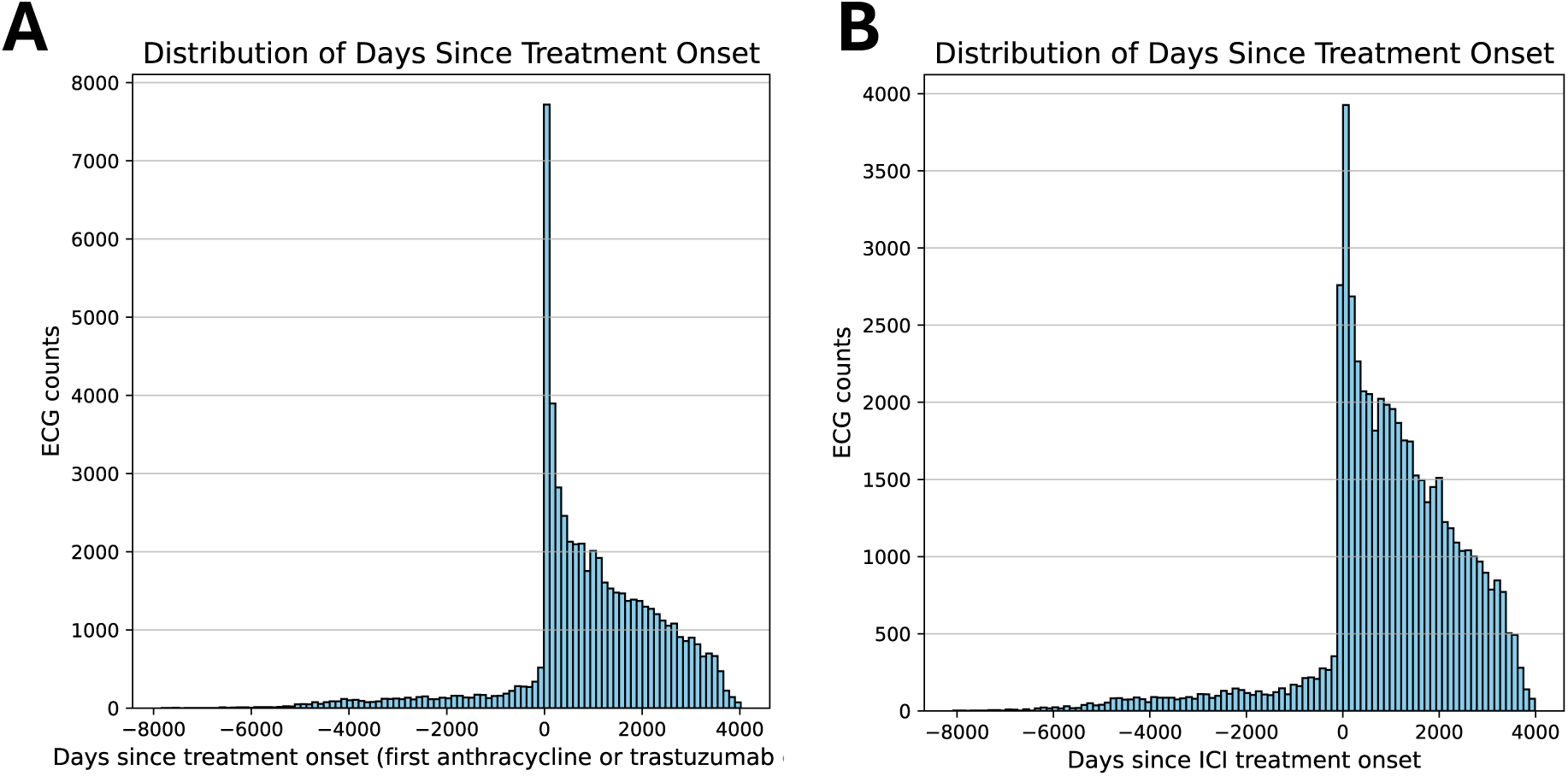
Timing of unique ECGs performed relative to the onset of treatment with anthracyclines and/or trastuzumab, or ICI. The plots show histograms of unique ECG counts with the x axis denoting the time difference between the first treatment cycle with anthracyclines and/or trastuzumab (A), and ICIs (B). Negative labels reflect ECGs performed before treatment was started. ECG: electrocardiograms; ICI: immune checkpoint inhibitors.

**Figure 3.**
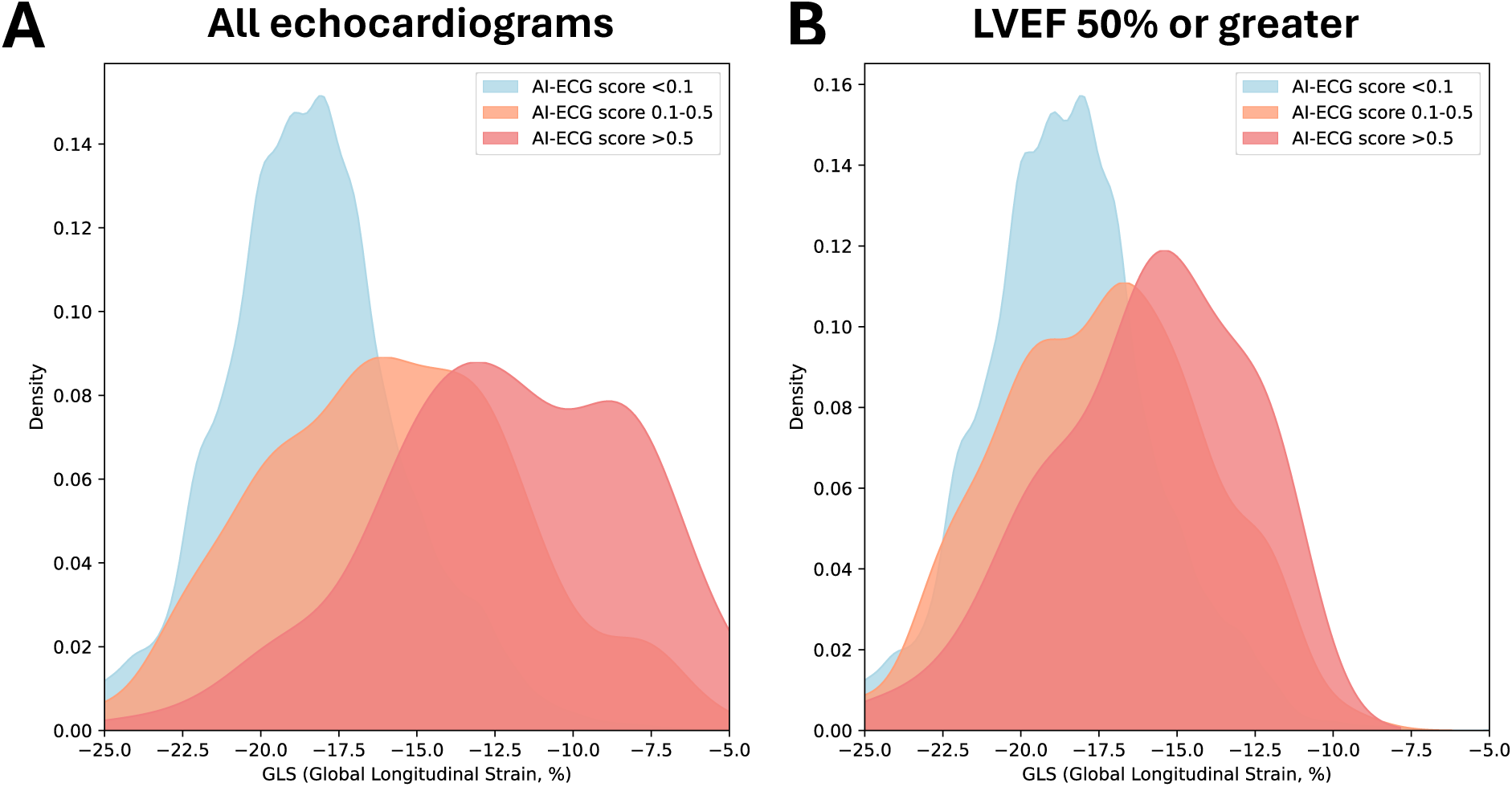
Distribution of global longitudinal strain (GLS) across AI-ECG LVSD groups. Density plots summarizing GLS across subgroups of AI-ECG LVSD scores (<0.1, 0.1 to <0.5, and 0.5 or greater) for all pairs of ECGs-echocardiograms performed within 15 days of each other (**A,** *n*=6333, 759 and 179, respectively), as well as in a subgroup of echocardiograms with reported LVEF of 50% or greater (**B,** *n*=6049, 536, and 60, respectively). AI: artificial intelligence; ECG: electrocardiography; GLS: global longitudinal strain; LVSD: left ventricular dysfunction.

### AI-ECG and future CTRCD with anthracyclines or trastuzumab

We identified a total of 1,308 patients (age 59 [IQR 49-67] years, 999 [76.4%] women) with breast cancer (n=801, 61.2%) and/or non-Hodgkin lymphoma (n=560, 42.8%) who had available *baseline* 12-lead ECG within a median of 1 day before [IQR: 29 days before to 1 day after] treatment initiation, and no prior history of cardiomyopathy. More than two in three (n=952, 72.8%) received treatment with doxorubicin, and one in four (n=336, 25.7%) treatment with trastuzumab (**Table 1)**. The median AI-ECG LVSD prediction at baseline was 0.01 [IQR 0.00-0.02], with 81 (6.2%) participants screening positive (≥0.1) for LVSD at baseline. Over a median follow-up of 80 [IQR 42-115] months, there were 404 (30.9%) cases meeting our CTRCD criteria, and 327 (25.0%) deaths. Higher baseline AI-ECG scores were significantly and independently associated with a higher risk of the composite definition of CTRCD (adj. HR 2.22 [95%CI: 1.63-3.02], *p*<0.001) (**Figure 4a**). In a secondary analysis of LVEF-based outcomes, positive AI-ECG screening was associated with a 3.7-fold higher risk of developing an LVEF<50% (n=122 events, HR_adjusted_ 3.66 [95%CI: 2.25-5.94], *p*<0.001), and a 4.8-fold higher risk of developing an LVEF<40% (n=72 events, HR_adjusted_ 4.76 [95%CI: 2.62-8.66], *p*<0.001) (**Figure 4b**). Higher AI-ECG LVSD scores were also predictive of worse overall survival (n=327 deaths, HR_adjusted_ 1.78 [95%CI: 1.26-2.52], *p*<0.001).

**Figure 4.**
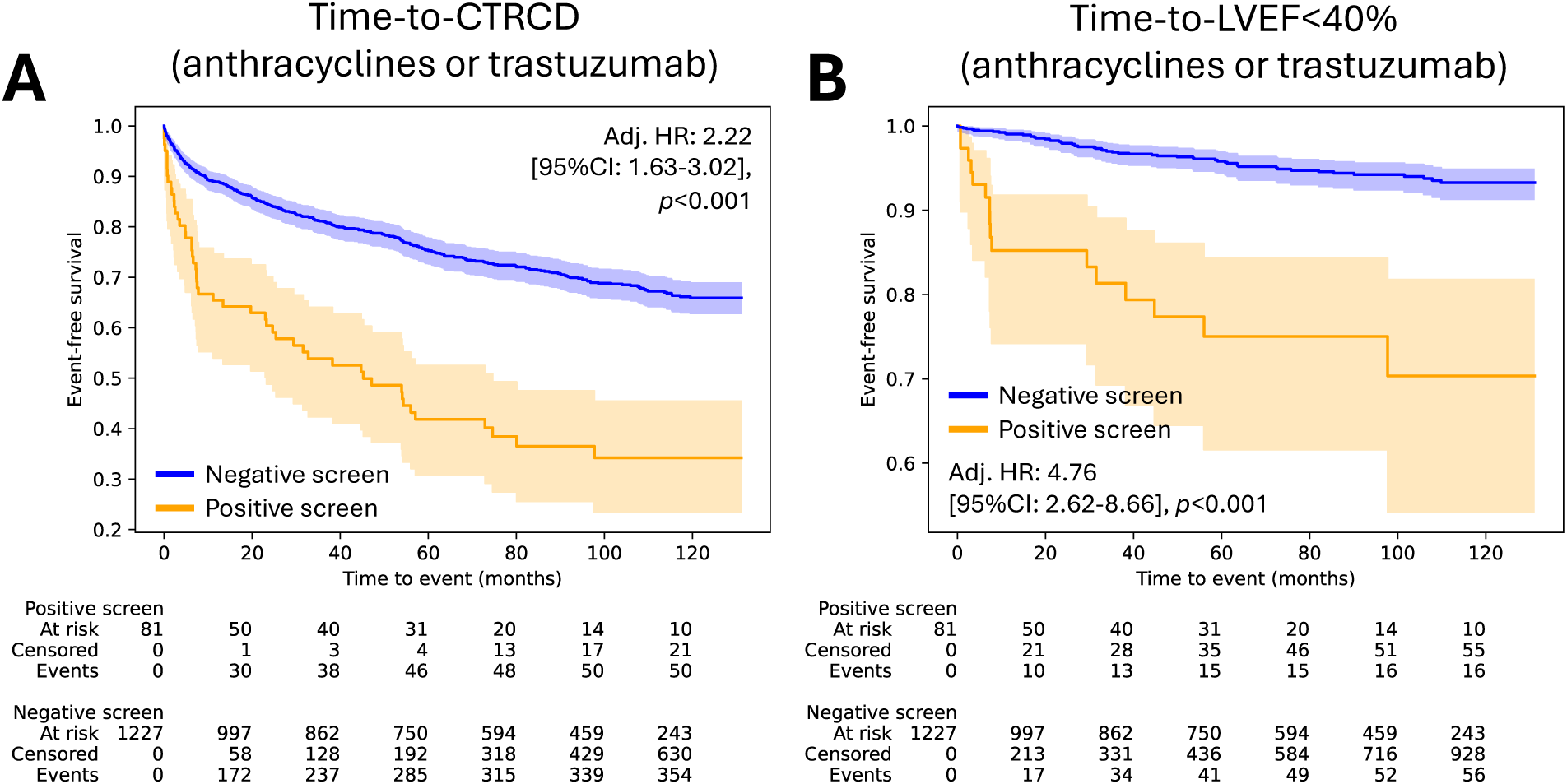
Baseline AI-ECG screening for LVSD and incidence of CTRCD among patients receiving anthracyclines/trastuzumab therapy. Kaplan-Meier survival curves for event-free survival and adjusted Cox regression-derived hazard estimates for CTRCD (defined as any LVEF<50%, cardiomyopathy or heart failure) **(A)** or LVEF<40% **(B)** stratified by positive versus negative screening by AI-ECG at baseline. AI: artificial intelligence; CI: confidence interval; CTRCD: Cancer-therapeutic related cardiac dysfunction; ECG: electrocardiography; HR: hazard ratio; LVEF: left ventricular ejection fraction; LVSD: left ventricular dysfunction.

### Dynamic changes in AI-ECG LVSD predictions prior to CTRCD

When further analyzing the trends in the AI-ECG scores in 12-month intervals leading up to the time of CTRCD (or the last known follow-up for those who did not experience the event), we observed a progressive rise in the AI-ECG scores of the CTRCD group relative to controls, with significant differences noted as early as 12-to-24 months before diagnosis (**Figure 5**). In **Figure 6**, we provide a representative example of a person in their 80s who developed CTRCD after initiating treatment with an anthracycline-based regimen. The initial ECGs were consistent with negative screens (AI-ECG LVSD predictions of 0.001 and 0.014 within 3 and ∼6 months of treatment initiation). However, a 12-lead ECG performed approximately 12 months later demonstrated new conduction and ST/T abnormalities, with the AI-ECG model now providing a positive screen with a probability of 0.998, shortly before an echocardiogram confirmed a new reduction in LVEF to <20% from >55% at baseline.

**Figure 5.**
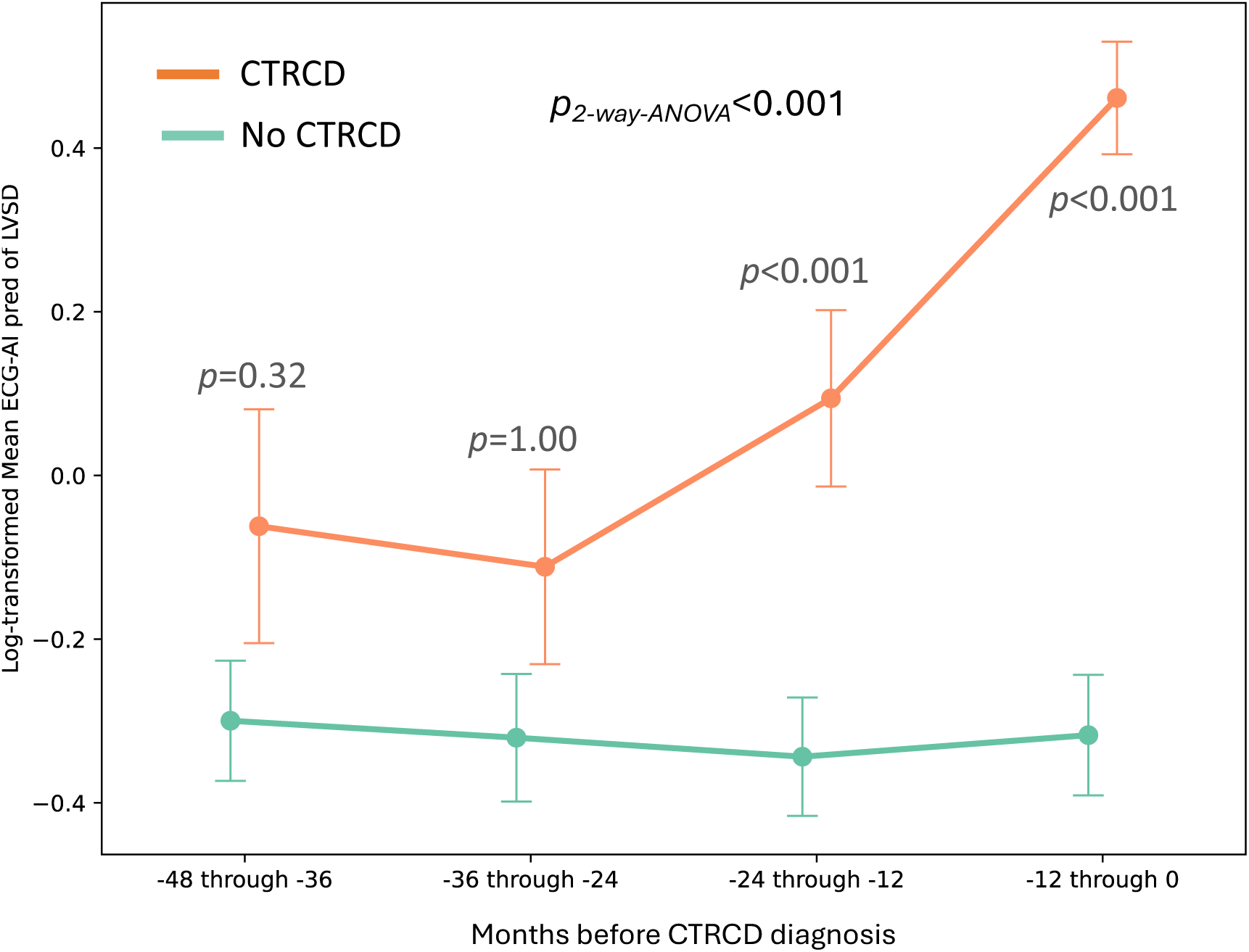
Evolution in AI-ECG LVSD predictions before CTRCD diagnosis. Means and standard errors of mean (SEM) for the log-transformed AI-ECG LVSD predictions in 12-month intervals leading up to the date of CTRCD diagnosis (or last known follow-up for patients without CTRCD). The comparison of AI-ECG LVSD predictions across groups of patients with CTRCD (orange) vs those without (green) and time was performed using 2-way analysis of variance (ANOVA). Post-hoc comparisons were performed by unpaired t-tests with correction for multiple comparisons using the Bonferroni method. AI: artificial intelligence; CTRCD: Cancer-therapeutic related cardiac dysfunction; ECG: electrocardiography; LVSD: left ventricular dysfunction.

**Figure 6.**
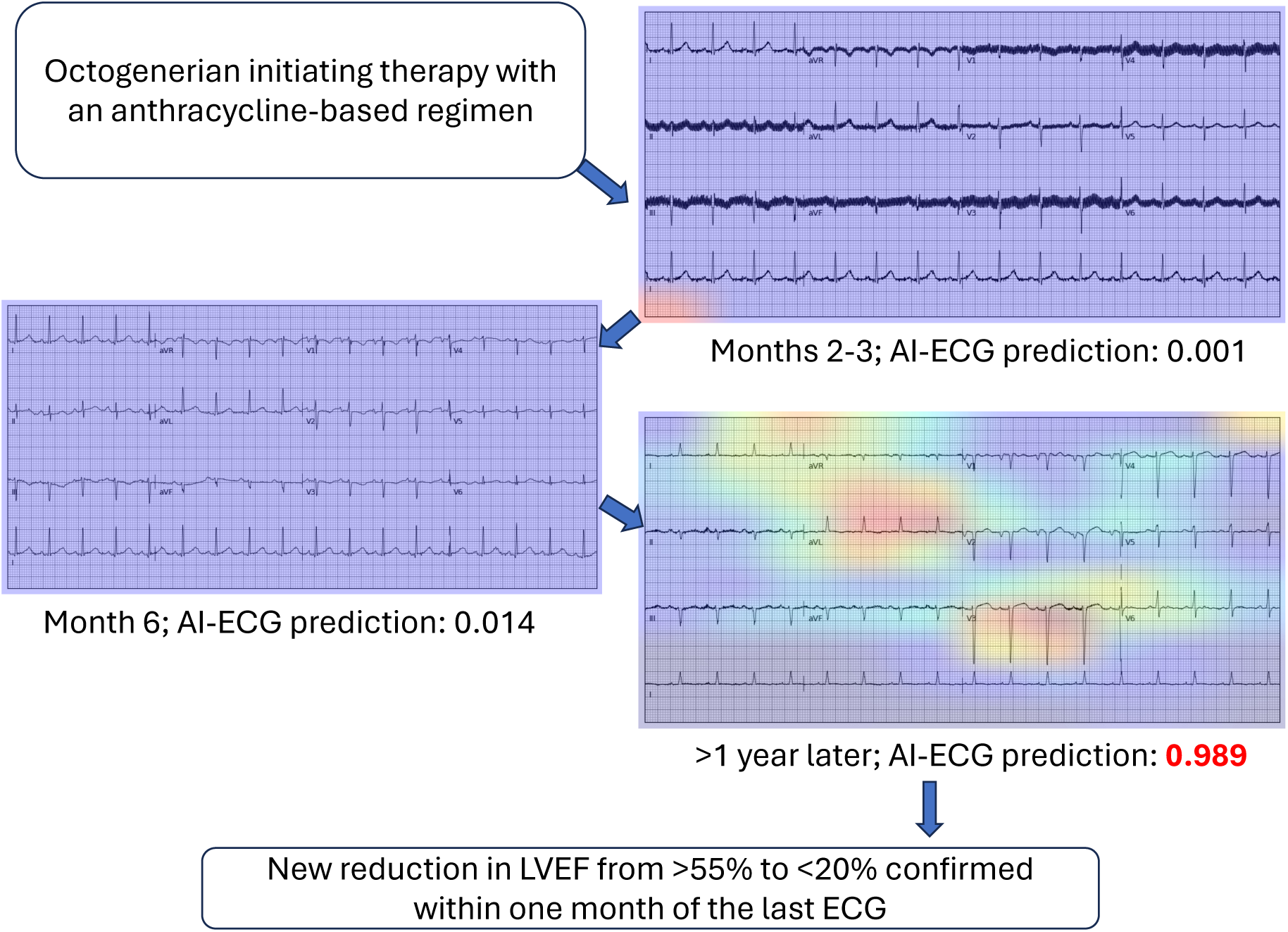
An example of longitudinal changes in AI-ECG LVSD predictions before CTRCD diagnosis. An example of an octogenarian treated with an anthracycline-based regimen. The figure shows the longitudinal evolution in the 12-lead ECG appearance and the GradCAM (activation/saliency maps) of the AI-ECG LVSD predictions across time, leading up to the diagnosis of CTRCD more than a year following treatment initiation, confirmed by an echocardiographic decline in LVEF to <20% from >55% at baseline. GradCAM visualization of a 12-lead ECG image shortly before the echocardiographic confirmation shows the development of ST-T changes, predominantly in lead V3 and aVL, which largely drive the model’s predictions. AI: artificial intelligence; CTRCD: cancer therapeutics-related cardiac dysfunction; ECG: electrocardiography; GradCAM: Gradient-weighted Class Activation; LVEF: left ventricular ejection fraction; LVSD: left ventricular systolic dysfunction.

### AI-ECG and future myocarditis in patients undergoing treatment with ICI

We identified 2,056 patients who received ICI (age 65 [IQR 57-73] years, 913 (44.4%) women) and had available baseline 12-lead ECG within a median of −1 [-19 to 0] days before treatment initiation, with no prior history of cardiomyopathy. The most common agent was pembrolizumab (n=1055, 51.3%), whereas the most common cancer diagnosis was lung cancer (n=978, 47.6%), followed by melanoma (n=364, 17.7%). The median AI-ECG LVSD prediction was 0.01 [IQR 0.00-0.03], with 204 (9.9%) of participants screening positive (≥0.1 for LVSD at baseline). Over a median follow-up of 63 [IQR 28-99] months, there were 35 (1.7%) myocarditis cases, and 965 (46.9%) deaths. Baseline AI-ECG scores were not associated with the risk of downstream myocarditis (HR_adjusted_ 1.36 [0.47-3.93], *p*=0.57) (**Figure 7**), or death (HR_adjusted_ 1.15 [0.95-1.41], *p*=0.16) in this population.

**Figure 7.**
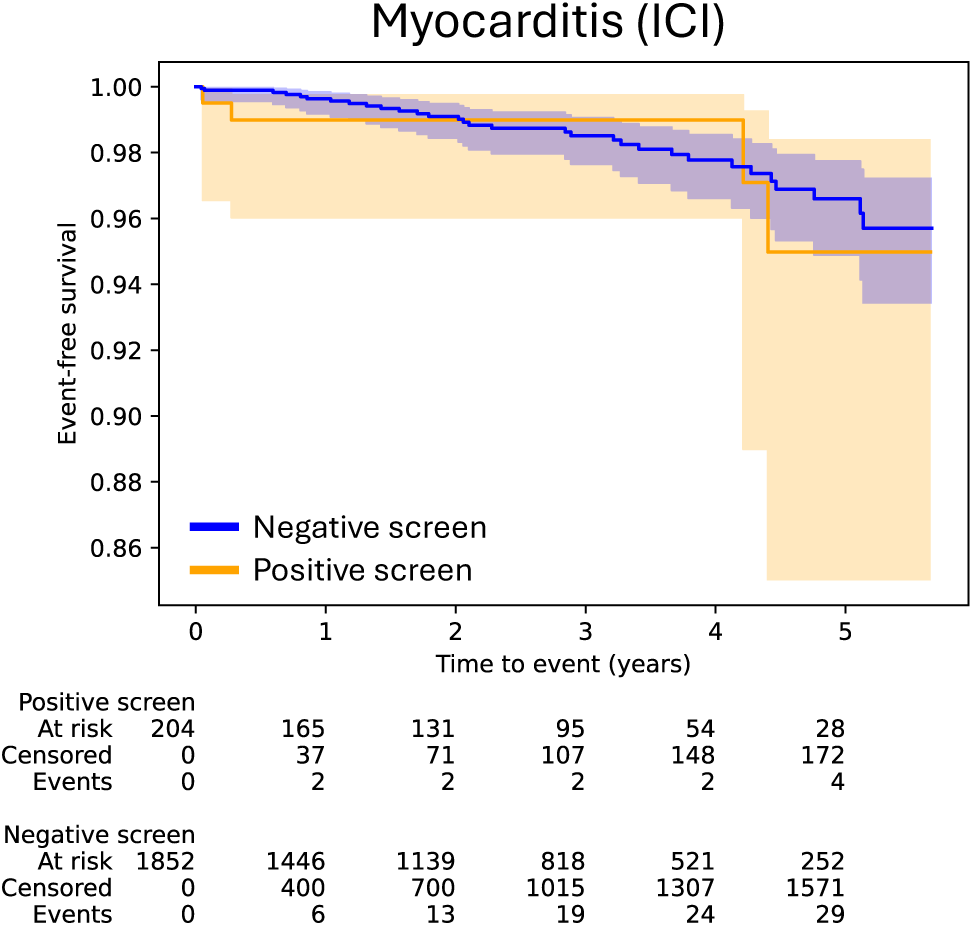
Baseline AI-ECG screening for LVSD and incidence of myocarditis among patients receiving ICI therapy. Kaplan-Meier survival curves for event-free survival and adjusted Cox regression-derived hazard estimates for myocarditis stratified by positive versus negative screening by AI-ECG at baseline. AI: artificial intelligence; CI: confidence interval; CTRCD: Cancer-therapeutic related cardiac dysfunction; ECG: electrocardiography; HR: hazard ratio; ICI: immune checkpoint inhibitors.

## DISCUSSION

We demonstrate a validated AI-ECG biomarker that only requires images of ECGs and can identify individuals with normal systolic function at baseline and no prior cardiomyopathy who are at increased risk of developing CTRCD following exposure to anthracyclines or trastuzumab. A positive AI-ECG screen at baseline translated into a nearly 2 to 5-fold higher adjusted risk of developing new cardiomyopathy or heart failure during follow-up, independent of the baseline risk profile. Further to the prognostic value of baseline AI-ECG, we provide evidence that, across the spectrum of cancer-related cardiotoxicity, AI-ECG predictions exhibit a dynamic course, with a significant increase relative to controls as early as 12-24 months before the clinical diagnosis of anthracycline or trastuzumab-related cardiomyopathy. Collectively, these findings support the use of a validated, accessible, and scalable technology as an additional read-out in guiding the risk stratification of patients referred for potential cardiotoxic chemotherapy.

Our findings supplement a rapidly expanding body of literature in cardio-oncology, as summarized in recently published guidelines and clinical consensus statements.^3, 9^ Given its ubiquitous nature and low cost, ECG is widely endorsed as a class I recommendation for the baseline cardiovascular risk stratification before initiating potentially cardiotoxic chemotherapy, including anthracyclines and/or trastuzumab.^3, 15^ Transthoracic echocardiography (TTE),^31^ including strain imaging by GLS,^17, 18^ and circulating biomarker characterization with natriuretic peptide and troponin measurements,^3, 31–34^ are also routinely recommended according to the risk profile of the proposed regimen and each individual patient. More recently, multimodal imaging with magnetic resonance imaging, including T1 mapping, extracellular volume (ECV) quantification,^11, 14^ or positron emission tomography with radiotracers tracking myocardial function, metabolism, and tissue repair have also shown value in pre-clinical or small clinical studies.^14, 35, 36^ However, access to these technologies varies across different parts of the world,^37–39^ underlining an unmet need for efficient cardiovascular risk stratification that can be performed at the point-of-care using readily accessible, yet personalized technologies.

Our work builds on recent developments in the AI-ECG space,^12, 13, 26, 40–42^ highlighting the ability of convolutional neural networks and other deep learning architectures to extract features that identify hidden and previously unrecognized labels of cardiovascular disease. However, it is innovative in several key aspects. First, our model, which is freely available online and directly accessible to any user,^28^ enables direct screening for a label of cardiac function that has traditionally required echocardiography, cardiovascular magnetic resonance or nuclear multi-gated acquisition scans for its accurate quantification.^43^ This provides an inexpensive, yet globally available technology for the monitoring and risk stratification of some of the most common forms of cancer and widely used agents in oncology. Second, unlike previously described and validated algorithms that rely on ECG signals,^12, 42^ our approach may use any real-world ECG image as input. This enables scalable inference at the point-of-care without requiring local health systems to reconfigure their electronic health record pipelines or establish such systems where they are not available. This directly addresses one of the key goals of medical AI, which is to build solutions that attenuate rather than exacerbate existing disparities in healthcare access.^44^ Finally, our work expands recent reports on the use of signal-based AI-ECG models as diagnostic tools to detect concomitant LVEF reduction detectable on echocardiography, as shown in an analysis of 683 female breast cancer patients who were treated with anthracycline chemotherapy, with an area under the receiver operating characteristic (AUROC) curve of 0.93 for detecting concomitant LVEF of less than 50%.^12^ Critically, the present work expands this to baseline risk stratification at treatment initiation, supporting the prognostic and diagnostic value of AI-ECG detection of LVSD. Furthermore, we provide data supporting that AI-ECG predictions may dynamically evolve over the course of the treatment, potentially identifying at risk individuals as early as 12-24 months before clinical diagnosis, findings that stand consistent with longitudinal cohort studies using multimodal imaging.^45–47^

### Limitations

Certain limitations merit consideration. First, our study was a retrospective cohort analysis including patients and monitoring practices that predated the most recent cardio-oncology guidelines. As a result, there was variation in the intensity and frequency of cardiovascular monitoring and diagnostic testing. This further limited our ability to integrate circulating biomarkers as additional readouts or covariates in our models. We feel, however, that the present study lays a solid foundation for a prospective clinical trial of ambulatory AI-ECG-enabled monitoring. Second, there are numerous combinations of potentially cardiotoxic agents and cancer types and subtypes, all with different baseline risk profiles and comorbidities, which were impossible to model in the context of this study. Third, we did not account for concomitant medication changes, or additional cancer-directed therapies, such as radiation therapy, which could potentially impact the cardiovascular profile of these patients. Since such interventions may occur anytime during follow-up or before the start date of these agents, we opted for a descriptive, observational analysis of our cohort and did not model for these time-varying effects.

## CONCLUSION

An AI tool that takes ECG images as input offers a robust and scalable solution to identify individuals at high risk of developing CTRCD in response to cardiotoxic chemotherapy. Our findings support the potential utility of AI-ECG monitoring for the cost-effective risk stratification and triage of patients receiving anthracyclines or anti-HER-2 therapies.

## PERSPECTIVES

### Core Clinical Competencies

- Cancer therapeutics may exert cardiotoxic effects that limit their effectiveness. There is an unmet need to efficiently stratify the risk of cancer therapeutics-related cardiac dysfunction (CTRCD) before treatment initiation using personalized approaches.

### Translational Outlook Implications

- Artificial intelligence (AI) applied to electrocardiographic (ECG) images can identify individuals with a 2-to-5-fold higher adjusted risk of developing CTRCD following exposure to anthracyclines or trastuzumab.
- AI-ECG predictions exhibit dynamic trends, with an increase in predictions detected as early as 12 to 24 months before the clinical diagnosis of CTRCD.

## Data Availability

Data produced in the present study are available upon reasonable request to the authors.

## ABBREVIATIONS LIST

AC: anthracyclines
AI: artificial intelligence
CCS(R): Clinical Classification Software (Refined)
CI: confidence interval
CTRCD: cancer therapeutics-related cardiac dysfunction
ECG: electrocardiography
EHR: electronic health record
GLS: global longitudinal strain
HER2: human epidermal growth factor receptor-2 (HER2) receptor inhibitors
HR: hazard ratio
ICD: International Classification of Diseases
ICI: immune checkpoint inhibitors
IQR: interquartile range
LVSD: left ventricular systolic dysfunction.

## Notes

### Competing Interest Statement

R.K. is an Associate Editor of JAMA and receives research support, through Yale, from the Blavatnik Foundation, Bristol-Myers Squibb, Novo Nordisk, and BridgeBio. He is a coinventor of U.S. Provisional Patent Applications 63/177,117, 63/428,569, 63/346,610, 63/484,426, 63/508,315, 63/580,137, 63/606,203, 63/562,335, and a co-founder of Ensight-AI, Inc and Evidence2Health, LLC. E.K.O. is an academic co-founder of Evidence2Health LLC, and an ad hoc consultant for Caristo Diagnostics, Ltd. He is a co-inventor in patent applications (US17/720,068, 63/619,241, 63/177,117, 63/580,137, 63/606,203, 63/562,335, WO2018078395A1, WO2020058713A1) and has received royalty fees from technology licensed through the University of Oxford, outside the submitted work. VS is a coinventor of 63/346, 610 and 63/484,426, and a co-founder of Ensight-AI, Inc. H.M.K. works under contract with the Centers for Medicare & Medicaid Services to support quality measurement programs, was a recipient of a research grant from Johnson & Johnson, through Yale University, to support clinical trial data sharing; was a recipient of a research agreement, through Yale University, from the Shenzhen Center for Health Information for work to advance intelligent disease prevention and health promotion; collaborates with the National Center for Cardiovascular Diseases in Beijing; receives payment from the Arnold & Porter Law Firm for work related to the Sanofi clopidogrel litigation, from the Martin Baughman Law Firm for work related to the Cook Celect IVC filter litigation, and from the Siegfried and Jensen Law Firm for work related to Vioxx litigation; chairs a Cardiac Scientific Advisory Board for UnitedHealth; was a member of the IBM Watson Health Life Sciences Board; is a member of the Advisory Board for Element Science, the Advisory Board for Facebook, and the Physician Advisory Board for Aetna; and is the co-founder of Hugo Health, a personal health information platform, and co-founder of Refactor Health, a healthcare AI-augmented data management company, and Ensight-AI, Inc. All other authors declare no competing interests.

### Funding Statement

National Heart, Lung, and Blood Institute of the National Institutes of Health (under awards R01HL167858 and K23HL153775 to RK, and 1F32HL170592-01 to EKO).

### Summary of Updates

We have updated the funding statement to acknowledge funding through the National Institutes of Health (NIH)/NHLBI. The title has also been revised to better reflect the content of the study.

