## Supplementary material for "Artificial intelligence-enhanced risk stratification of cancer therapeutics-related cardiac dysfunction using electrocardiographic images": Online Supplement

**Online-only Supplement**

Evangelos K. Oikonomou MD DPhil[a], Veer Sangha BS[a,b], Lovedeep S. Dhingra MBBS[a], Arya Aminorroaya MD MPH[a], Andreas Coppi PhD[c], Harlan M. Krumholz MD SM[a,c], Lauren A. Baldassarre MD[a], Rohan Khera MD MS[a,c,d,e*]

[a] Section of Cardiovascular Medicine, Department of Internal Medicine, Yale School of Medicine, New Haven, CT, USA
[b] Department of Engineering Science, University of Oxford, Oxford, UK
[c] Center for Outcomes Research and Evaluation, Yale-New Haven Hospital, New Haven, CT, USA
[d] Section of Biomedical Informatics and Data Science, Yale School of Medicine, New Haven, CT, USA
[e] Section of Health Informatics, Department of Biostatistics, Yale School of Public Health, New Haven, CT, USA

**Brief title:** AI-ECG-guided monitoring of cancer therapy cardiotoxicity

[*]**Corresponding author:**
Rohan Khera, MD, MS
195 Church St, 6th Floor, New Haven, CT 06510
203-764-5885;; @rohan_khera

### Supplemental Tables

#### Online Table 1. Diagnosis codes used in the study.

| | ICD-9 starting with: | ICD-10 starting with: | CCS(R): |
|---|---|---|---|
| **Hypertension** | '401', '4010', '4011', '4019', '402', '4020', '4021', '4029', '403', '4030', '4031', '4039', '404', '4040', '4041', '4049', '6420', '6422', '6427', '6429' | 'I10', 'I11', 'I110', 'I119', 'I12', 'I120', 'I129', 'I13', 'I130', 'I131', 'I132', 'I139', 'I674', 'O10', 'O100', 'O101', 'O102', 'O103', 'O109', 'O11' | 'CIR007', 'CIR008' |
| **Diabetes mellitus** | '250', '2500', '25000', '25001', '25009', '2501', '25010', '25011', '25019', '2502', '25020', '25021', '25029', '2503', '2504', '2505', '2506', '2507', '2509', '25090', '25091', '25099', '6480' | 'E10', 'E100', 'E101', 'E102', 'E103', 'E104', 'E105', 'E106', 'E107', 'E108', 'E109', 'E11', 'E110', 'E111', 'E112', 'E113', 'E114', 'E115', 'E116', 'E117', 'E118', 'E119', 'E12', 'E120', 'E121', 'E122', 'E123', 'E124', 'E125', 'E126', 'E127', 'E128', 'E129', 'E13', 'E130', 'E131', 'E132', 'E133', 'E134', 'E135', 'E136', 'E137', 'E138', 'E139', 'E14', 'E140', 'E141', 'E142', 'E143', 'E144', 'E145', 'E146', 'E147', 'E148', 'E149', 'O240', 'O241', 'O242', 'O243', 'O249' | 'END003', 'END002' |
| **Cardiomyopathy or Heart failure** | "425", "4254", "4258", "4259", "428", "4280", "4281", "4282", "4283", "4284", "4289" | "I42", "I420", "I427", "I428", "I429", "I50", "I501", "I502", "I503", "I504", "I508", "I509" | “CIR005” |
| **Stroke** | '433', '4330', '4331', '4332', '4333', '4338', '4339', '434', '4340', '4341', '4349', '435', '4359', '437', '4370', '4371' | 'G45', 'G450', 'G451', 'G452', 'G453', 'G454', 'G458', 'G459', 'I63', 'I630', 'I631', 'I632', 'I633', 'I634', 'I635', 'I638', 'I639', 'I64','I65', 'I650', 'I651', 'I652', 'I653', 'I658', 'I659', 'I66', 'I660', 'I661', 'I662', 'I663', 'I664', 'I668', 'I669', 'I672', 'I693', 'I694' | "CIR020", "CIR022", "CIR025" |
| **Ischemic Heart Disease** | '410', '4109', '411', '4119', '412', '4129', '413', '4139', '414', '4140', '4148', '4149' | 'I20', 'I200', 'I208', 'I209', 'I21', 'I210', 'I211', 'I212', 'I213', 'I214', 'I219', 'I21X', 'I22', 'I220', 'I221', 'I228', 'I229', 'I23', 'I230', 'I231', 'I232', 'I233', 'I234', 'I235', 'I236', 'I238', 'I24', 'I240', 'I241', 'I248', 'I249', 'I25', 'I250', 'I251', 'I252', 'I255', 'I256', 'I258', 'I259', 'Z951', 'Z955' | "CIR009", "CIR011" |
| **Peripheral arterial disease** | '4402', '4442' | 'I702', 'I7020', 'I7021', 'I742', 'I743', 'I744' | "CIR023", "CIR026" |
| **Chronic Kidney Disease** | '403', '4030', '4031', '4039', '404', '4040', '4041', '4049', '585', '5859', '6421', '6462' | 'I12', 'I120', 'I13', 'I130', 'I131', 'I132', 'I139', 'N18', 'N180', 'N181', 'N182', 'N183', 'N184', 'N185', 'N188', 'N189', 'Z49', 'Z490', 'Z491', 'Z492' | "GEN003" |
| **Myocarditis** | “422”, “4220”, “4229”, “4290” | “I40”, “I41”, “I514” | - |

CCS(R): Clinical Classification Software (Refined); ICD: International Classification of Diseases.

### Online Table 2. Procedural diagnosis codes used.

| | CPT codes: |
|---|---|
| **Coronary Artery Bypass Grafting (CABG)** | "33510", "33511", "33512", "33513", "33514", "33515", "33516", "33517", "33518", "33519", "33521", "33522", "33523", "33533", "33534", "33535", "33536" |
| **Percutaneous coronary intervention (PCI)** | "92920", "92921", "92922", "92923", "92924", "92928", "92929", "92933", "92934", "92937", "92938", "92941", "92943", "92944" |
| **Peripheral arterial disease intervention** | "27590", "27591", "27592", "27598", "27880", "27881", "27882", "35452", "35454", "35456", "35459", "35470", "35472", "35473", "35474", "35521", "35533", "35541", "35546", "35548", "35549", "35551", "35556", "35558", "35563", "35565", "35566", "35571", "35582", "35583", "35585", "35587", "35621", "35623", "35641", "35646", "35651", "35654", "35656", "35661", "35663", "35665", "35666", "35671", "37205", "37206", "37207", "37208" |

CPT: Current Procedural Terminology.
